# The use of social media and online dating among individuals with unipolar depression and bipolar disorder

**DOI:** 10.1101/2020.05.20.20102707

**Authors:** Klara F. K. Rydahl, René B. K. Brund, Clara R. Medici, Vibeke Hansen, Krista M. N. Straarup, Sune P. V. Straszek, Søren D. Østergaard

## Abstract

**Background:** Studies on how individuals with affective disorders use and perceive their use of social media are lacking from the literature.

**Methods:** A questionnaire focusing on affective disorders and the use of social media and online dating was handed out to outpatients with unipolar depression or bipolar disorder and general practice patients with or without affective disorders (latter as controls). The association between affective disorders and the use of social media and online dating was visualized using network graphs and analyzed using linear/logistic regression.

**Results:** A total of 194 individuals with unipolar depression, 124 individuals with bipolar disorder and 196 controls were included in the analysis. Having unipolar depression or bipolar disorder was not associated with the time spent on social media compared with controls. Using the controls as reference, having bipolar disorder was associated with use of online dating (adjusted odds ratio: 2.2 (95% CI: 1.3; 3.7)). The use of social media and online dating had a mood-congruent pattern with decreased and more passive use during depressive episodes, and increased and more active use during hypomanic/manic episodes. Among the respondents with affective disorder, 51% reported that social media use had an aggravating effect on symptoms during mood episodes, while 10% reported a beneficial effect. For online dating, the equivalent proportions were 49% (aggravation) and 20% (benefit), respectively.

**Limitations:** Selection and report biases may have affected the results of the survey.

**Conclusions:** The use of social media and online dating seems related to symptom deterioration among individuals with affective disorders.

## Introduction

With its rapid growth in both number of users (Ortiz-Ospina, 2019) and time spent on use (Clement, 2019; OECD, 2019), social media platforms like Facebook, Instagram and Twitter have become virtually ubiquitous, and profoundly changed the way humans interact (Ortiz-Ospina, 2019). This development has spurred a growing research field investigating how this new way of interacting affects the mental well-being of the users. The majority of these studies suggest an association between social media use and lower mental well-being, lowered self-esteem, and increased symptoms of anxiety and depression (Chou and Edge, 2012; Hanna et al., 2017; McCrae et al., 2017; Riehm et al., 2019; Shakya and Christakis, 2017; Shaw et al., 2015; Tromholt, 2016; Vannucci et al., 2017; Yoon et al., 2019). While causality is inherently difficult to establish in this case, studies taking temporality into account (investigating mental health downstream of social media use (Shakya and Christakis, 2017)) as well as a randomized (though unblinded) intervention study (Tromholt, 2016) have generally confirmed the notion of social media use reducing mental well-being.

Interestingly, it seems that it is not only the quantity of social media usage that is responsible for the likely negative effect on mental health, but also the nature of this use (Aalbers et al., 2019; Gerson et al., 2017; Thorisdottir et al., 2019; Verduyn et al., 2015). Specifically, when dividing the use of social media into “active use” (text messaging, posting pictures, contributing to public debates etc.) and “passive use” (scrolling through feeds and watching other users’ updates, pictures and videos) (Gerson et al., 2017), there is some evidence to support that passive use may be more harmful than active use (Thorisdottir et al., 2019). This resonates well with the “upward social comparison” hypothesis (Boers et al., 2019; Hanna et al., 2017), which is based on the assumption that people tend to selectively display the most positive aspects of their lives on social media (active use) (Chou and Edge, 2012; Nadkarni and Hofmann, 2012) and that other people – who tend to take these positively biased projections at face value (during passive use) – therefore get the impression that their own life compares negatively to that of other social media users (Østergaard, 2017).

As outlined above, there is a growing body of evidence suggesting, that social media use may be harmful to mental health. This raises the question as to how individuals with established mental disorders use social media and how this use interacts with their illness. Given the negative cognitive bias characterizing the depressive phases of affective disorders (Kilford et al., 2015) and hence the likely hypersensitivity to upward social comparison, it seems particularly prudent to study social media use in this population. Therefore, we conducted a survey with three main objectives:

I. To study the use of social media among individuals with affective disorders compared with individuals without affective disorders – having emphasis on i) the time spent on social media, ii) the nature of use (active versus passive), and iii) potential changes in use during depressive and hypomanic/manic episodes, respectively.
II. To study the use of a particular type of social media – namely online dating (e.g. Tinder, Happn and Grindr) among individuals with affective disorders compared with individuals without affective disorders. This objective was included due to the pleasure-seeking and hypersexual behavior commonly seen in relation to manic episodes in bipolar disorder (Correll et al., 2014; Kopeykina et al., 2016; Wang et al., 2020), where the constant availability of online dating may represent a challenge.
III. To study the self-perceived impact of social media and online dating on the symptoms/course of affective episodes among individuals with affective disorders.

## Methods

### Design

We conducted a questionnaire-based survey, aimed at three different populations: individuals with unipolar depression, individuals with bipolar disorder and individuals without affective disorders (controls). The questionnaire was in Danish and made specifically for the purpose (an English translation of the questionnaire is included in the supplementary material). The survey was conducted in Denmark at four sites: 1) The outpatient unit for unipolar depression (including the “collaborative care” unit, where patients with depression are referred to by their general practitioner) at Aarhus University Hospital – Psychiatry, 2) The outpatient unit for bipolar disorder at Aarhus University Hospital – Psychiatry 3) an urban general practice, and 4) a rural general practice. All Danish citizens have free access to a general practitioner (and secondary/tertiary health care services), so the two general practices provided access to individuals from the general population without affective disorders, which served as controls in the study. An urban and a rural general practice were chosen deliberately to roughly reflect the catchment area of the outpatient units at Aarhus University Hospital – Psychiatry. Only individuals aged 18–75, who were current or prior users of social media and/or online dating, were invited to participate.

### Recruitment

The survey was conducted from October 2018 to May 2019 (except for the rural general practice where the recruitment ended in February 2019). The recruitment of participants was organized by the individual sites in order to fit the local logistics/clinical practices. At the rural general practice (one doctor practicing), the questionnaire was handed out directly to all patients meeting the age criterion. In the urban general practice (several doctors practicing), the questionnaire was available in the waiting room. At the outpatient units for unipolar depression and bipolar disorder, respectively, individual treatment providers (doctors, nurses and psychologist) invited their patients to participate on the basis of feasibility. This approach was required by the treatment providers to ensure that vulnerable patients with unipolar depression or bipolar disorder could discuss the questionnaire and/or their (emotional) response with a professional if needed. The participants’ answers to the questionnaire were however not disclosed to the doctors/treatment providers. This was specifically stated on the front page of the questionnaire.

### Questionnaire

The questionnaire consisted of three sections: Part I focused on socio-demographic and clinical aspects such as age, sex, educational level, relationship status and history of affective disorders (e.g. onset and number of depressive episodes, onset and number of manic/hypomanic episodes, and onset and number of mixed episodes). Part II focused on use of social media (e.g. time spent on social media, the use of passive and active functions, and use during depressive, and hypomanic/manic episodes), while part III focused on the use of online dating (questions analogue to part II).

### Ethics

The study was registered with the Danish Data Protection Agency and the data were processed and stored in accordance with the European Union General Data Protection Regulation. The Danish National Ethics Committee waives the requirement for ethical oversight for questionnaire-based surveys in accordance with the Law on Ethics Committees §14, 2: http://www.nvk.dk/forsker/naar-du-anmelder/hvilke-projekter-skal-jeg-anmelde anmelder/hvilke-projekter-skal-jeg-anmelde. On the front page of the questionnaire, it was stated that handing in the completed questionnaire to the doctor/treatment provider would be considered as consent for use of the data for research purposes.

### Definition of respondents with unipolar depression, bipolar disorder and controls

The respondents were divided into three groups (unipolar depression, bipolar disorder and controls) based on the study site they were recruited from as well as their answers to the questionnaire:

#### Respondents from the two psychiatric outpatient units

As a general rule, the respondents from the two outpatient units (for unipolar depression and for bipolar disorder) were grouped according to the unit they attended. However, there were some exceptions to this rule. As individuals with bipolar disorder are not treated at the outpatient unit for unipolar depression and individuals with unipolar depression are not treated at the outpatient unit for bipolar disorder, we excluded “misclassified” individuals (n = 16), i.e. if there was a mismatch between their reported psychopathology and the unit they attended. A detailed explanation of this procedure is provided in the supplementary material.

#### Respondents from the two general practices

These respondents were categorized according to their answers to the questions regarding affective psychopathology:

#### Unipolar depression

Those reporting a history of one or more depressive episodes (and no hypomanic/manic/mixed episodes) and/or gave consistent answers to questions regarding social media and online dating use during depressive episodes (question number 22–25 and/or number 47 and 48).

#### Bipolar disorder

Those reporting a history of either a manic, hypomanic or mixed episodes and/or providing consistent answers to the questions regarding social media and online dating use during hypomanic/manic or mixed episodes (question number 26–36 and/or number 49–57).

#### Control group

if reporting neither hypomanic/manic, mixed nor depressive episodes.

### Exclusion

Respondents were excluded from the analyses for three reasons: I) respondents reporting no current or prior use of neither social media nor online dating, as they failed to meet the inclusion criterion for the study. II) Respondents who failed to report age or sex were excluded from the analyses as these variables were essential for the analyses. III) Respondents who were misclassified (see above as well as supplementary material).

### Handling of ambiguous responses

As the questionnaire was completed using pen-and-paper, some respondents occasionally put checks between boxes. This was handled by rounding off (away from zero). Specifically, for “Yes/No” response categories, a check between the boxes was classified as “yes”. Similarly, for a check between likert-scale response categories such as “No difference” and “Reduced/relieved to a mild degree”, the latter was chosen as the response used in the analyses.

### Merging of response categories

In order to have sufficient numbers to allow for statistical analyses, we merged likert-scale response categories as follows: For questions with five response categories including a neutral middle category, the two response categories on each side were merged, creating a three-level variable (−1, 0, +1). For questions with three response categories (e.g., “No”, “Yes, sometimes”, “Yes, often”), the two affirmative response categories were merged, creating a dichotomous variable.

### Statistical analyses

The three populations (unipolar depression, bipolar disorder and controls) were characterized using descriptive statistics. The association between unipolar depression/bipolar disorder and the use of social media and online dating was investigated by means of logistic and linear regression analyses, adjusted for age and sex (as well as relationship status in the case of online dating), as appropriate for the investigated outcome.

Furthermore, we visualized the pattern of use of social media and the motivation for using online dating by means of network analysis. Specifically, data regarding social media (stratified a priori into passive and active functions) and online dating use was plotted visually with the Fruchterman-Reingold algorithm (Fruchterman and Reingold, 1991) using the qgraph package (Epskamp et al., 2012). This algorithm presents strongly connected nodes (social media and online dating functions) central on the network graph and visualizes the degree of combined use of two functions/motivations via the thickness of edges. Each combination between functions of use/motivations were counted for each person and summarized in a symmetric matrix. The networks were unweighted and plots of the same questionnaire were averaged to produce network graphs that were comparable between populations. The edges were not penalized, such that use of two functions/motivations endorsed by at least one respondent is displayed as an edge with the lowest edge thickness, while progressively increased edge thickness indicates increasing numbers of respondents reporting use of specific functions/motivations.

### Post hoc analysis

Based on inspection of the network graphs illustrating the motivations for using online dating, we conducted a follow-up analysis of the association between “getting approval” and “finding a sexual partner” in order to determine whether this combination was more commonly endorsed among those with bipolar disorder compared with the controls. This analysis was carried out using logistic regression adjusted for age, sex and relationship status.

### Sensitivity analysis

The logistic regression analyses were evaluated for appropriate fit via the link test developed by Pregibon and influential observations were diagnosed by Pearson’s residual, deviance residual and Pregibon’s leverage, respectively (Pregibon, 1981). In cases where the link test indicated that the model was misspecified, we changed the right hand side of the link function based on hypotheses and visual inspection of different residual and leverage plots until the model was appropriately fitted. In cases of doubt regarding influential observations, we compared a model without these observations with the model with these observations. All linear regressions were evaluated by plotting the Pearson’s residuals to assess outliers and kernel density. P-P and Q-Q plots were used to evaluate the normal distribution of the residuals. In case of outliers, these were evaluated for influence on the estimates, while non-normal data was fitted by changing the function on the right hand side. Moreover, both the logistic and linear regressions were bootstrapped to assess the robustness of the estimates.

### Software

Data analysis was conducted in STATA 16 (StataCorp LP, College Station, TX) or R version 3.6.2 (R Core Team (2013)).

## Results

### Respondents

A total of 577 individuals from the four study sites responded to the survey (138 from the outpatient unit for unipolar depression, 134 from the outpatient unit for bipolar disorder, 116 from the rural general practice and 189 from the urban general practice). The questionnaires returned by 63 of the respondents were not included in the analyses for the following reasons: I) eight respondents reported no current/prior use of social media or online dating, II) 39 respondents did not include information on either sex and/or age, and III) 16 respondents were misclassified (see the Supplementary Material). Of the remaining 514 respondents that constituted the final sample for analysis, 194 had a history of unipolar depression (116 from the outpatient unit and 78 from the general practitioners), 124 had a history of bipolar disorder (118 from the outpatient unit and 6 from the general practitioners), and 196 had neither unipolar depression nor bipolar disorder (controls). A flowchart illustrating the recruitment and the definition of the final sample for analysis is available in the Supplementary Material.

Table 1 shows data regarding demographics, episodes of unipolar depression/bipolar disorder, and use of social media/online dating for the 514 respondents included in the analyses. Of these, 513 used social media, 266 used online dating, and 265 used both social media and online dating. The majority (74%) of the respondents were female – relatively equally distributed across the three groups. The controls were older, more likely to be married, have children, and to have a higher education compared with the respondents with unipolar depression and bipolar disorder.

**Table 1:** Demographic, clinical and social media/online dating characteristics of the sample.

|  | Unipolar depression | Bipolar disorder | Controls |
| --- | --- | --- | --- |
|  | n=194<br>37.8% | n= 124<br>24.1% | n=196<br>38.1% |
| **Sex, n(%)** |  |  |  |
| Female | 145 (74.7) | 98 (79) | 140 (71.4) |
| **Age, median, (Q: 25% ;75%)** |  |  |  |
|  | 30 (24;42) | 30 (25;40) | 36 (25;53) |
| **Place of Birth, n (%)** |  |  |  |
| Denmark | 183 (94.3) | 115 (92.7) | 187 (95.4) |
| Outside Denmark | 11 (5.7) | 9 (7.3) | 9 (4.6) |
| **Marital Status, n (%)** |  |  |  |
| Single | 77 (39.7) | 50 (40.3) | 33 (16.8) |
| Partner, lives alone | 26 (13.4) | 22 (17.8) | 18 (9.2) |
| Living with a partner | 44 (22.7) | 30 (24.2) | 54 (27.6) |
| Married | 42 (21.6) | 18 (14.5) | 80 (40.8) |
| Divorced/separated | 5 (2.6) | 4 (3.3) | 11 (5.6) |
| **Children, n (%)** |  |  |  |
| Yes | 85 (43.8) | 44 (35.5) | 110 (56.1) |
| No | 109 (56.2) | 80 (64.5) | 86 (43.9) |
| **Education level, n (%)** |  |  |  |
| Primary & secondary school | 27 (14) | 15 (12.1) | 13 (6.6) |
| Upper Secondary School | 54 (27.8) | 41 (33.1) | 42 (21.4) |
| Vocational Education | 7 (3.6) | 6 (4.8) | 17 (8.7) |
| Short-cycle higher education | 32 (16.5) | 9 (7.3) | 27 (13.8) |
| Medium-cycle higher education | 53 (27.3) | 36 (29) | 52 (26.5) |
| Long cycle higher education | 21 (10.8) | 17 (13.7) | 45 (23) |
| **Age at onset of depression, n (%)** |  |  |  |
| < 18 years old | 31 (16.3) | 44 (36.7) |  |
| 18-24 years old | 84 (44.2) | 46 (38.3) |  |
| 25-34 years old | 39 (20.5) | 16 (13.3) |  |
| ≥35 years old | 36 (19) | 14 (11.7) |  |
| Respondents, n | 190 | 120 |  |
| **Number of depressive episodes, n (%)** |  |  |  |
| 1 episode | 102(54) | 14 (11.8) |  |
| 2-5 episodes | 69 (36.5) | 57 (47.9) |  |
| 6-10 episodes | 7 (3.7) | 22 (18.5) |  |
| ≥ 11 episodes | 11 (5.8) | 26 (21.8) |  |
| Respondents, n | 189 | 119 |  |
| **Age at onset of hypomanic/ manic episodes, n (%)** |  |  |  |
| < 18 years old |  | 5 (4.6) |  |
| 18-24 years old |  | 50 (45.9) |  |
| 25-34 years old |  | 30 (27.5) |  |
| ≥35 years old |  | 24 (22) |  |
| Respondents, n |  | 109 |  |
| **Number of manic episodes, n (%)** |  |  |  |
| 1 episode |  | 31 (28.7) |  |
| 2-5 episodes |  | 44 (40.7) |  |
| 6-10 episodes |  | 14 (13) |  |
| ≥ 11 episodes |  | 19 (17.6) |  |
| Respondents, n |  | 108 |  |
| **Age at onset of mixed episodes, n (%)** |  |  |  |
| < 18 years old |  | 5 (5.2) |  |
| 18-24 years old |  | 45 (46.9) |  |
| 25-34 years old |  | 24 (25.0) |  |
| ≥/ >35 years old |  | 22 (22.9) |  |
| Respondents, n |  | 96 |  |
| **Number of mixed episodes, n (%)** |  |  |  |
| 1 episode |  | 24 (25.8) |  |
| 2-5 episodes |  | 33 (35.5) |  |
| 6-10 episodes |  | 9 (9.7) |  |
| ≥ 11 episodes |  | 27 (29.0) |  |
| Respondents, n |  | 93 |  |
| **Use of social media** |  |  |  |
| **Social media users, n (%)** |  |  |  |
| Social media user (current or prior use) | 194 (100) | 124 (100) | 195 (99.5)<sup>a</sup> |
| Respondents, n | 194 | 124 | 195 |
| **Time spent on social media, median, (Q: 25%;75%)** |  |  |  |
| Minutes per day | 90 (60;180) | 90 (60;180) | 68 (45;120) |
| Respondents, n | 189 | 122 | 191 |
| **Social media account created, n (%)** |  |  |  |
| 0-5 years ago | 13 (6.7) | 7(5.7) | 34 (17.4) |
| 6-8 years ago | 45 (23.2) | 27 (21.7) | 53 (27.2) |
| ≥ 9 years ago | 136 (70.1) | 90 (72.6) | 108 (55.4) |
| Respondents, n | 194 | 124 | 195 |
| **Use of online dating** |  |  |  |
| **Online dating users, n (%)** |  |  |  |
| Online dating user (current or prior use) | 103 (53.1) | 84 (67.7) | 79 (40.3) |
| Respondents, n | 188 | 120 | 194 |
| **Time spent on online dating, median, (Q: 25%;75%)** |  |  |  |
| Minutes per day | 60 (19;90) | 60 (29;120) | 30 (20;60) |
| Respondents, n | 102 | 78 | 77 |
| **Online dating account created, n (%)** |  |  |  |
| < 1 year ago | 7 (6.8) | 6 (7.2) | 4 (5.1) |
| 1-2 years ago | 12 (11.7) | 15 (18.1) | 11 (13.9) |
| 3-5 years ago | 44 (42.7) | 27 (32.5) | 33 (41.8) |
| 6-8 years ago | 18 (17.5) | 16 (19.3) | 12 (15.2) |
| ≥ 9 years ago | 22 (21.4) | 19 (22.9) | 19 (24.1) |
| Respondents, n | 103 | 83 | 79 |

<sup>a</sup> one individual who used online dating did not use (other) social media platforms

### Social media use among individuals with unipolar depression and bipolar disorder

The median time spent on social media (daily average use from first account was activated) was 60 minutes among respondents with unipolar depression as well as those with bipolar disorder, and 30 minutes for the controls. However, following adjustment for age and sex, neither unipolar depression nor bipolar disorder was associated with increased time spent on social media at the level of statistical significance (see Table 2A).

**Table 2:** The association between affective disorders and use of social media (A) and online dating (B)

| Time spent on social media with and without adjustment for age and sex<sup>a</sup> |  |  |  |
| --- | --- | --- | --- |
|  | Crude coefficient (95% CI) | Mutually adjusted (95% CI) | P-value |
| Unipolar depression<sup>b</sup> | 29.3 (11.1; 47.6) | 16.9 (-0.5; 34.2) | 0.058 |
| Bipolar disorder<sup>b</sup> | 35.0 (14.3; 55.6) | 18.1 (-1.7; 37.9) | 0.073 |
| Age |  | -2.3 (-2.9; -1.8) |  |
| Sex<sup>c</sup> |  | -10.2 (-27.4; 7) |  |

**B.**
| Online dating use (yes/no) with and without adjustment for age and sex<sup>a</sup> |  |  |  |
| --- | --- | --- | --- |
|  | Crude OR (95% CI) | Mutually adjusted (95% CI) | P-value |
| Unipolar depression<sup>b</sup> | 1.7 (1.1; 2.5) | 1.4 (0.9; 2.2) | 0.1 |
| Bipolar disorder<sup>b</sup> | 3.1 (2.0; 5) | 2.6 (1.6; 4.2) | **<0.001** |
| Age |  | 1.0 (0.9; 1.0) |  |
| Sex<sup>c</sup> |  | 1.2 (0.8; 1.9) |  |
| Online dating use (yes/no) with and without adjustment for age, sex and relationship status |  |  |  |
|  |  | Mutually adjusted OR (95% CI) | P value |
| Unipolar depression<sup>b</sup> |  | 1.2 (0.8; 1.9) | 0.392 |
| Bipolar disorder<sup>b</sup> |  | 2.2 (1.3; 3.7) | **0.002** |
| Age |  | 1.0 (0.9; 1.0) |  |
| Sex<sup>c</sup> |  | 1.1 (0.7; 1.8) |  |
| Relationship<sup>d</sup> |  | 0.4 (0.3; 0.6) |  |
| Time spent on online dating with and without adjustment for age and sex |  |  |  |
|  | Crude coefficient (95% CI) | Mutually adjusted (95% CI) | P value |
| Unipolar depression<sup>b</sup> | 3.4 (-22; 28.8) | 4.6 (-20.6; 29.8) | 0.721 |
| Bipolar disorder<sup>b</sup> | 41.5 (14.5; 68.5) | 43.0 (16.1; 69.9) | **0.002** |
| Age |  | 1.1 (0.1; 2.1) |  |
| Sex<sup>c</sup> |  | -11.1 (-34.4; 12.5) |  |
| Time spent on online dating adjusted for age, sex and relationship status |  |  |  |
|  |  | Mutually adjusted (95% CI) | P-value |
| Unipolar depression<sup>b</sup> |  | 9.9 (-15.1; 34.8) | 0.438 |
| Bipolar disorder<sup>b</sup> |  | 49.2 (21.5; 75.9) | **<0.001** |
| Age |  | 1.1 (0.0; 1.9) |  |
| Sex<sup>c</sup> |  | -9.2 (-32.2; 13.9) |  |
| Relationship<sup>d</sup> |  | 34.1 (13.3; 54.9) |  |

Time spent on social media and online dating was analyzed using linear regression analysis, while online dating use was analyzed using logistic regression analysis.
<sup>a</sup> Among those using social media/online dating, respectively.
<sup>b</sup> Reference: Controls
<sup>c</sup> Reference: Female
<sup>d</sup> Reference: In a relationship (Having a partner and married)

The network graphs in Figure 1 illustrate the use of 16 social media functions for the respondents with unipolar depression, bipolar disorder, and the controls, respectively. Generally, the most widely used functions are the “passive” ones in combination with mainly three “active” functions, namely “chatting with friends/family/acquaintances”, “posting photos” and “participating in “groups”“. There were no clear differences in the pattern of use between the three groups of respondents. When stratifying the controls (which were older than the members of the two other groups) into those aged ≤31 years and > 31 years (median split), we observed a tendency towards more use of picture posting, event creation and following of public figures among the younger users.

**Figure 1.**
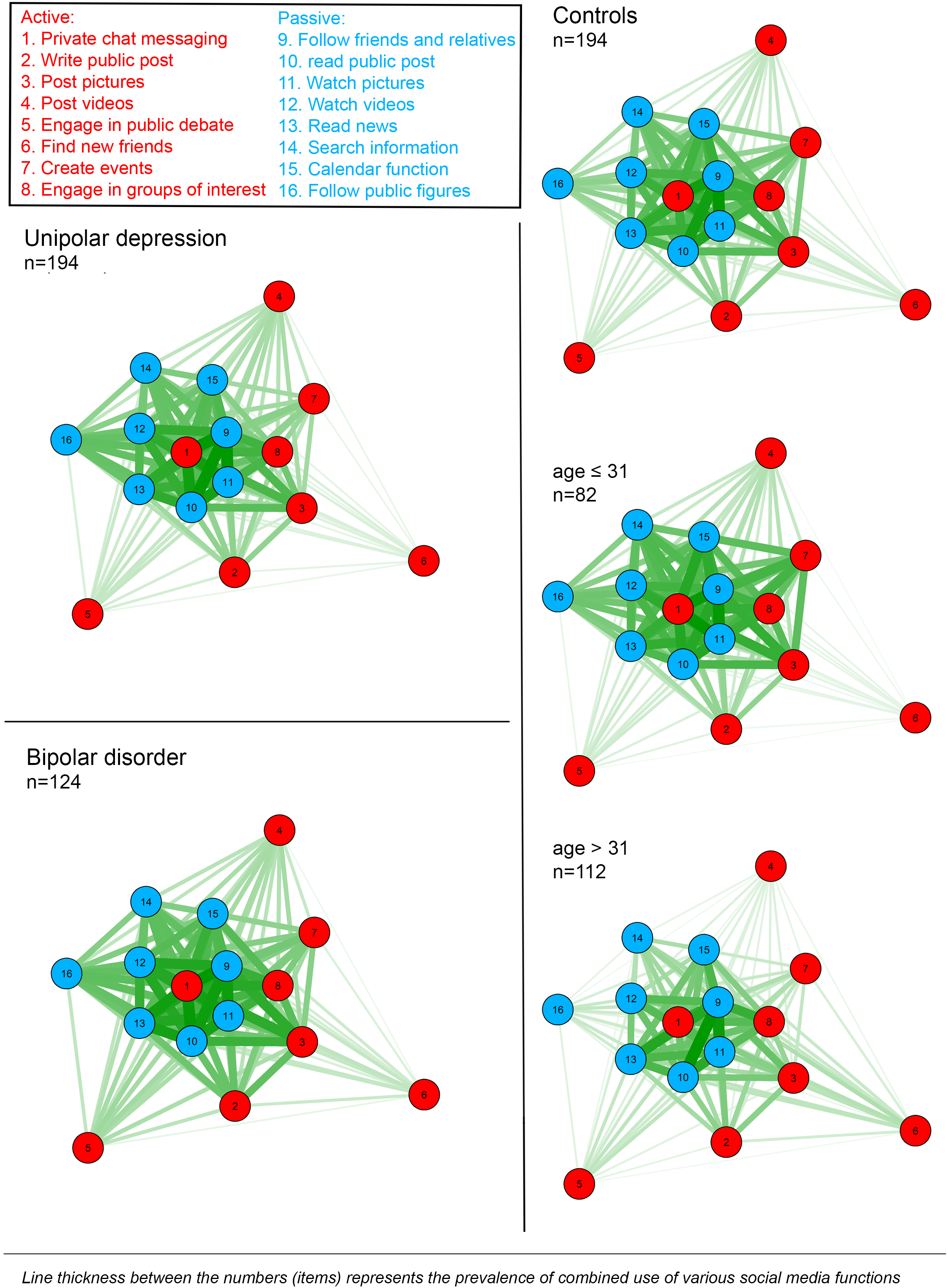
Network graphs showing the use of various social media functions.

### Changes in use of social media during depressive and hypomanic/manic episodes

Among the respondents with unipolar depression reporting use of social media, 90% responded to the questions concerning social media use during depressive episodes (questions 22–25). Among the respondents with bipolar disorder reporting use social media, 91% responded to the questions concerning social media use during depressive episodes, and 89% responded to the questions concerning use of social media during hypomanic/manic episodes (questions 26–36). Among the respondents with unipolar depression using social media during a depressive episode, 24% reported no change in social media activity, 30% reported increased activity, and 46% reported decreased activity. Similarly, among the respondents with bipolar disorder using social media during a depressive episode, 13% reported no change in social media activity, 29% reported increased activity, and 58% reported decreased activity. Among the respondents with bipolar disorder using social media during a hypomanic/manic episode, 9% reported no change in social media activity, 74% reported increased activity and 17% reported decreased activity.

Figure 2 illustrates how the use of social media functions changes during depression and hypomania/mania according to the respondents. Both the respondents with unipolar depression and bipolar disorder reported a strong tendency towards increased use of passive functions and decreased use of active functions during depressive episodes. During manic episodes, there appeared to be a more “global” increase/decrease across social media functions.

**Figure 2.**
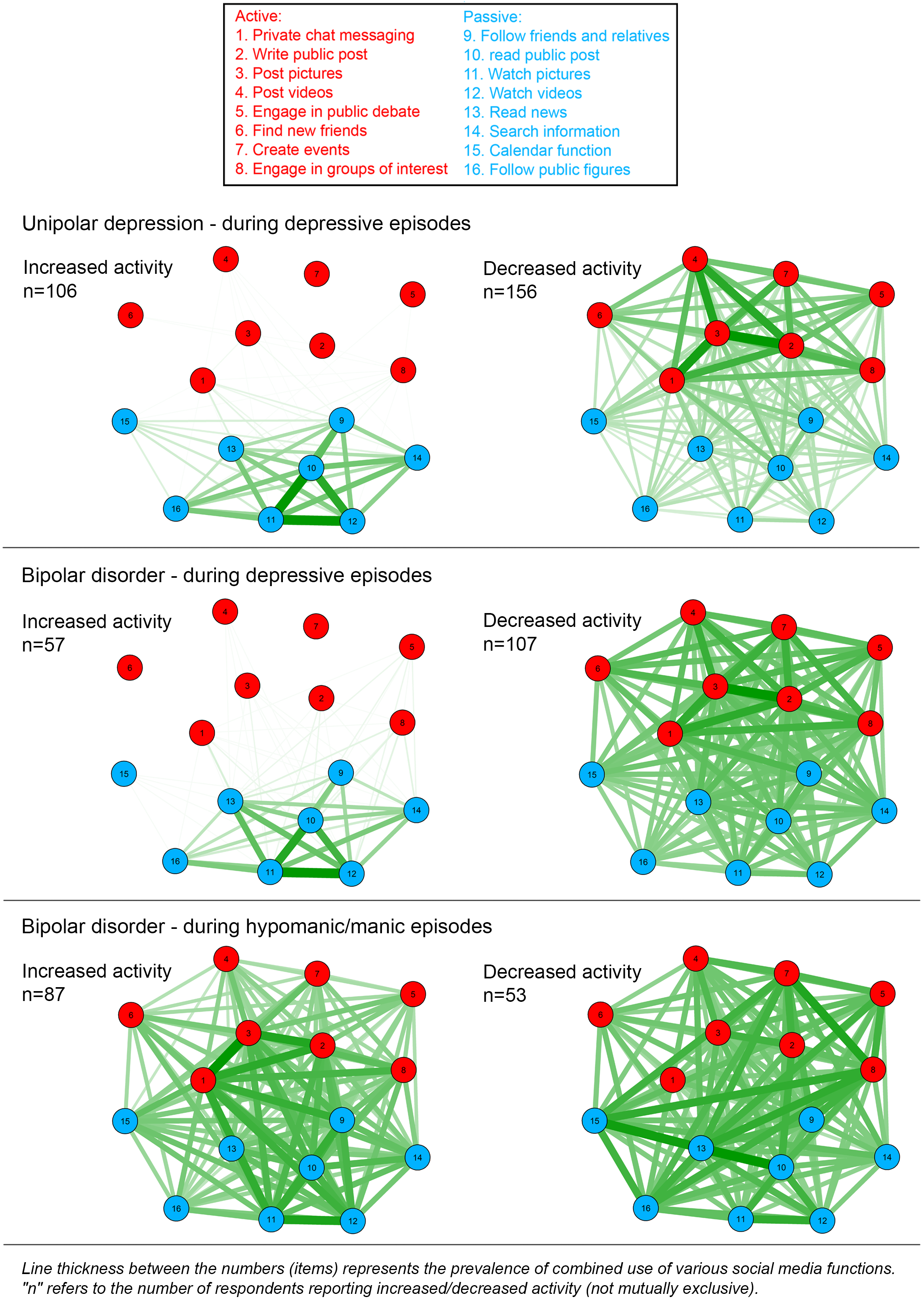
Network graphs showing change in use of social media functions during depressive and hypomanic/manic episodes.

### Use of online dating

The results of the analyses of the association between affective disorders and use of online dating are shown in Table 2B. Using the controls as reference, there was a statistically significant association between bipolar disorder and use of online dating (odds ratio (OR) adjusted for age and sex: 2.6 (95% CI: 1.6; 4.2)), which remained (OR: 2.2 (95% CI: 1.3; 3.7)) following additional adjustment for relationship status (dichotomously defined: Yes: Married, living together, having a partner but living alone. No: Single, divorced/separated, widow). Furthermore, among the respondents using online dating – using the controls as reference – there was a statistically significant association between bipolar disorder and spending more time on online dating (regression coefficient adjusted for age and sex: 43.0 (95% CI: 16.1; 69.9)), which also remained (regression coefficient: 49.2 (95% CI: 21.5; 75.9)) following additional adjustment for relationship status (defined as above).

The network graphs in Figure 3 illustrate the motivations for using online dating among the respondents with unipolar depression, bipolar disorder, and the controls, respectively. Across the three groups, the most frequent combinations of motivations were a) “finding a romantic partner” and “getting approval”, b) “finding a romantic partner” and “finding a sexual partner”, and c) “getting approval” and “finding a sexual partner”. When stratifying the controls (which were older than the members of the two other groups) into those aged ≤31 years and > 31 years (median split), it became evident that the “getting approval” and “finding a sexual partner” combination of motivations was more common among the users aged ≤31, while the “finding a romantic partner” and “finding new friends” combination was more common among those aged > 31. We noticed that the “getting approval” and “finding a sexual partner” combination appeared to be particularly common among the respondents with bipolar disorder using online dating. Therefore, we investigated this association in a post hoc logistic regression analysis, which showed that the respondents with bipolar disorder were indeed significantly more likely than the controls to endorse this combination (OR after adjustment for age and sex: 3.4 (95% CI: 1.5–7.8), and OR: 3.1 (95% CI: 1.3–7.1) following additional adjustment for relationship status.

**Figure 3.**
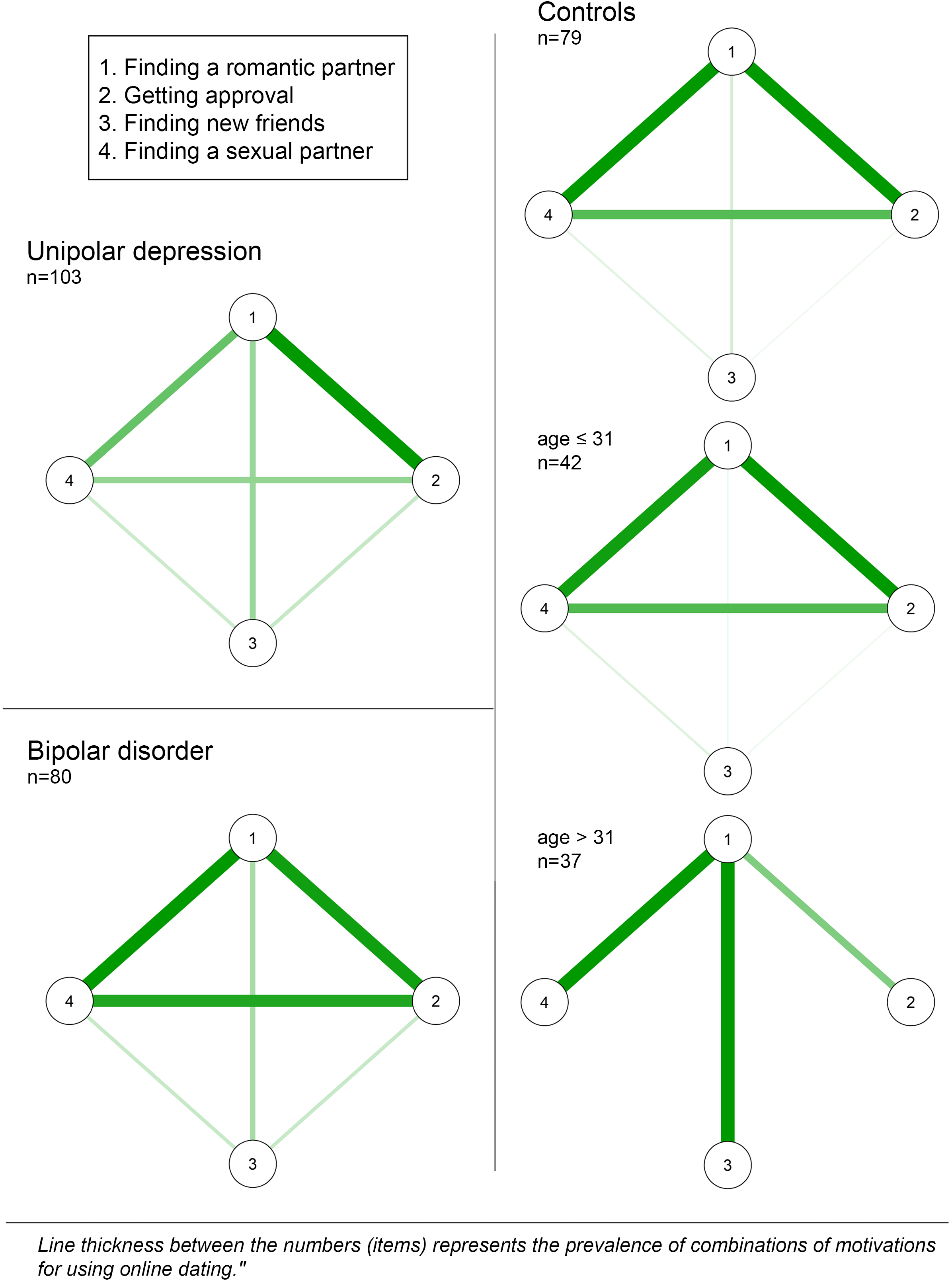
Network graphs showing the motivations for using online dating.

### Changes in use of online dating during depressive and hypomanic/manic episodes

Among the respondents with unipolar depression reporting use of online dating, 69% responded to the questions concerning use of online dating during depressive episodes (questions 47 and 48). Here, 27% reported that they experienced no change in the use of online dating during depressive episodes, 24% that they were more active, and 49% answered that they were less active during depressive episodes. Among the respondents with bipolar disorder reporting use of online dating, 69% responded to the questions concerning use of online dating during depressive episodes. Here, 7% reported that they experienced no change in the use of online dating during depressive episodes, 22% that they were more active, and 71% that they were less active during depressive episodes. Among the respondents reporting use of online dating with bipolar disorder, 80% responded to the questions concerning use of during hypomanic/manic episodes (questions 49–57). Here, 5% reported that they experienced no change in the use of online dating during hypomanic/manic episodes, 92% that they were more active, and 3% that they were less active during hypomanic/manic episodes.

### Self-reported effect of social media and online dating on symptoms of affective disorders

The results regarding self-reported effect of social media and online dating on symptoms of affective disorders during affective episodes are shown in Figure 4. The general pattern – across episodes of unipolar depression, bipolar depression and hypomania/mania – was that more respondents reported aggravating than beneficial effects of social media and online dating.

**Figure 4.**
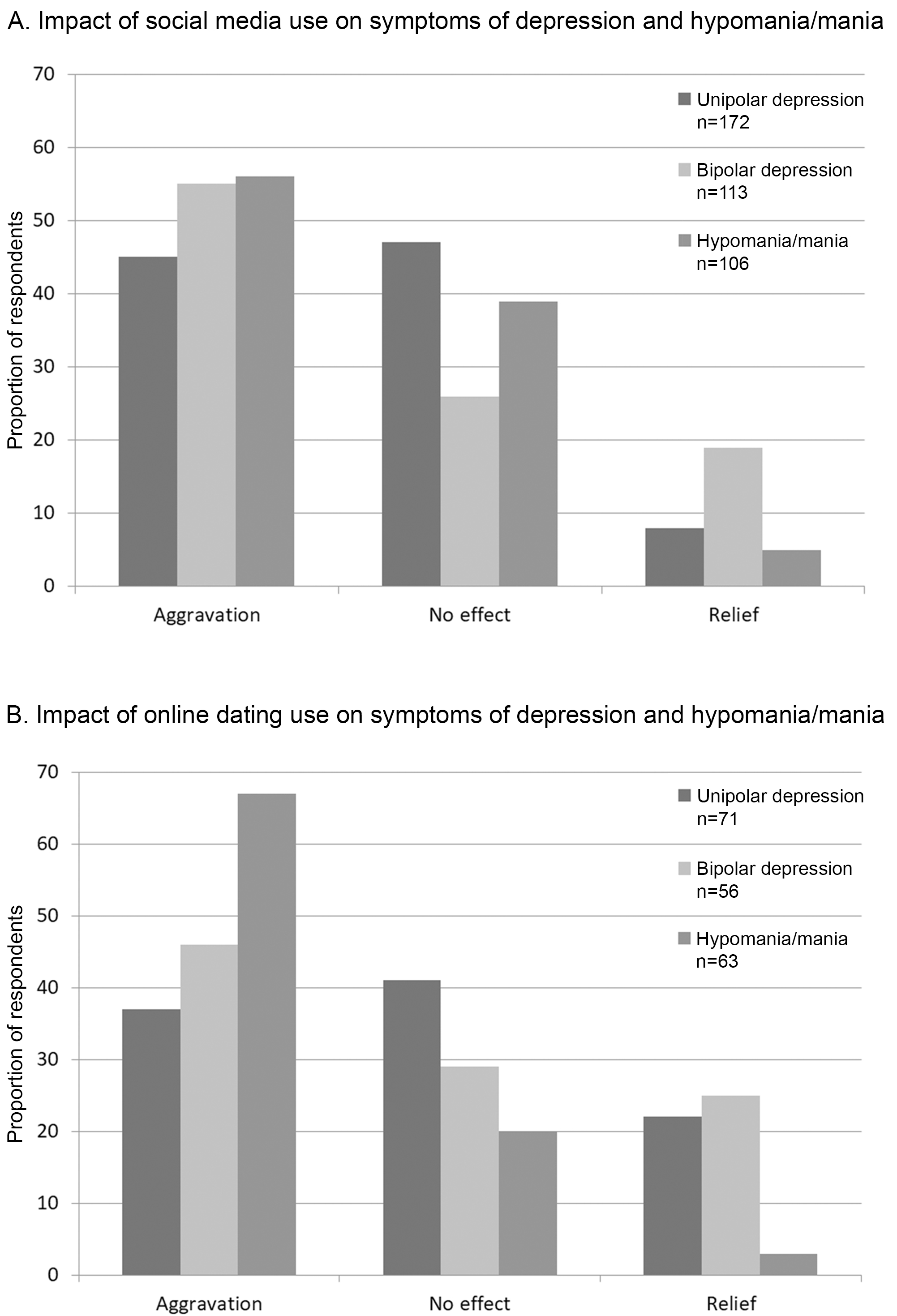
Self-reported effect of social media (A) and online dating use (B) on symptoms of affective disorders.

### Sensitivity analyses

The link test indicated that the logistic regression analyses were appropriately fitted and the residuals and leverage plots indicated no influential observations. Similarly, Pearson’s residuals, P-P and Q-Q plots showed that the assumptions were also met for the linear regression analyses. Similar differences in slope between groups was seen for the regression on time spent on social media and online dating when including or excluding potential influential observations (e.g. individuals spending more than 400 and 300 minutes and online dating sites each day, respectively) and confidence interval remained to cross zero.

## Discussion

The objective of this study was to explore the use of social media and online dating among individuals with unipolar depression and bipolar disorder as this aspect is of potential clinical importance due to reported associations between social media use and lower mental well-being, lowered self-esteem, and increased symptoms of anxiety and depression among general users of these platforms (Chou and Edge, 2012; Hanna et al., 2017; McCrae et al., 2017; Riehm et al., 2019; Shakya and Christakis, 2017; Shaw et al., 2015; Tromholt, 2016; Vannucci et al., 2017; Yoon et al., 2019). We focused on three aspects, namely I) The extent- and pattern of use of social media among individuals with unipolar depression and bipolar disorder – including potential changes in use during depressive and hypomanic/manic episodes, II) the extent of and motivation behind the use of online dating among individuals with unipolar depression and bipolar disorder – including potential changes in use during depressive and hypomanic/manic episodes, and III) the self-perceived impact of the use of social media and online dating on the symptoms of unipolar depression and bipolar disorder.

With regard to the use of social media, the time spent on this activity did not seem to differ between those with unipolar depression, bipolar disorder and the controls – when taking age and sex into account. As for the pattern of use, it was also largely similar across the groups, with a tendency towards more use of passive functions among the respondents with unipolar depression. While the overall use of social media among individuals with affective disorders appeared to be little different compared to that of controls, our findings point towards a marked mood-congruence in the use of social media among individuals with affective disorders. Specifically, the respondents reported a tendency towards decreased time spent on social media during both unipolar and bipolar depressive episodes – and increased time spent during hypomanic/manic episodes. Furthermore, the respondents reported that the use of passive functions increased, while the use of active functions decreased during both unipolar and bipolar depressive episodes. In contrast, during hypomanic/manic episodes, there was a tendency towards a global increase of all social media functions – including core active elements such as messaging and posting content. The apparent mood-congruent use of social media resonates well with the findings of Thorisdottir et al. (Thorisdottir et al., 2019), who found that passive social media use was associated with increased levels of anxiety and depressed mood, and that active social media use was associated with decreased levels of anxiety and depressed mood, among adolescents. However, due to the cross-sectional and non-experimental nature of both the present study and that of Thorisdottir et al., it is not possible to determine if this association is causal – nor the direction of potential causality. Nevertheless – from a clinical perspective – it seems somewhat problematic that the use of social media is predominantly passive during depression and active during hypomania/mania as this may contribute to sustaining these states of pathological mood/activity level (Lam et al., 2010, 2003; Scott et al., 2006).

The analysis focusing on online dating showed that individuals with bipolar disorder were more likely to engage in this activity. Furthermore, compared with the controls, who also used online dating, those with bipolar disorder spent more time on it, and were apparently more motivated by the perspective of getting approval and finding a sexual partner compared with the controls. This seems consistent with the tendency towards hypersexuality in bipolar disorder (Correll et al., 2014; Kopeykina et al., 2016; Wang et al., 2020). Finally, there was also a mood-congruent pattern in the use of online dating, with decreased time spent on this activity in relation to unipolar and bipolar depressive episodes, and increased time spent during hypomanic/manic episodes. As for the use of social media, this mood-congruence may be unfortunate with regard to sustaining depression and hypomania/mania (Lam et al., 2010, 2003; Scott et al., 2006).

In the questionnaire, the respondents were asked to evaluate the effect that the use of social media and online dating has upon their symptoms during depressive and hypomanic/manic episodes. The response to these questions was highly indicative of a negative effect of both social media and online dating use on the symptoms of depression and hypomania/mania. This could suggest that the concern raised above regarding the mood congruent use of social media and online dating is indeed relevant and should potentially be taken into consideration in clinical practice, where advice on balanced/healthy use of social media/online dating could easily be integrated in psychoeducation (Reinares et al., 2008; Soo et al., 2018; Tursi et al., 2013). Furthermore, it is worth mentioning that studies of social media use among individuals with other mental disorders, namely eating disorders (Mabe et al., 2014) and psychotic disorders (Berry et al., 2018), have also pointed towards mainly negative effects. With the tremendous role that social media is playing in today’s society, these results are worrying, and call for further studies investigating the effects of social media (including online dating) on mental health.

This study has three main limitations. First, relying on cross-sectional and self-reported data regarding the use of social media/online dating, as well its effect on symptoms of affective disorders is clearly not optimal (Boase and Ling, 2013; Deng et al., 2019; Junco, 2013; Scharkow, 2016). However, this project was unfortunately not logistically nor financially scaled to enable objective measures of both exposure and outcome over an extended period time from participants recruited at multiple sites. While it is inherently difficult to determine if and how this limitation has affected the results, it seems likely that the negative cognitive bias associated with depressed mood (Beck, 2008; Jabben et al., 2012; Kilford et al., 2015) and the positive cognitive bias associated with hypomania/mania (Kærsgaard et al., 2018; Schönfelder et al., 2017) may have contributed to the reported mood-congruence in the use of social media and online dating. For this reason, we suggest that future studies should ideally be based on a more sophisticated technical setup to enable more accurate and unbiased measures of both exposures and outcomes over an extended period of time. Second, the fact that the questionnaires were distributed differently at the four sites may have resulted in selection bias, where the two most obvious sources seem to be the following: i) In the urban general practice where the questionnaire was available in the waiting room, those without affective disorders who chose to participate may have a social media/online dating use that is not representative for the general population, and ii) At the outpatient units for unipolar depression and bipolar disorder, the treatment-providers handed out the questionnaire, and may – consciously or subconsciously – have done this selectively. The existence and impact of these biases cannot be determined based on the data at hand. However, future studies should aim at a more systematic and uniform recruitment strategy. Third and finally, social desirability bias may have affected the response to the more sensitive questions from the survey, for instance those relating to the motivation for using online dating (e.g. “Getting approval” and “Finding a sexual partner”) (Krumpal, 2013). The positive cognitive bias associated with hypomania/mania may be part of the reason why these motivations tended to be endorsed more often by the respondents with bipolar disorder.

To our knowledge, this study is among the very first to investigate the use of social media and online dating among individuals with unipolar depression and bipolar disorder. The main findings were the tendency towards mood-congruent use of social media and online dating (less/passive use during depression and more/active use during hypomania/mania), and the predominantly negative self-reported effects of social media and online dating use upon symptoms of depression and hypomania/mania (worsening). While these findings should ideally be replicated in other settings, it seems worth considering whether advice on balanced/healthy use of social media/online dating should be included in the psychoeducation provided in relation to treatment of unipolar depression and bipolar disorder.

## Data Availability

In agreement with Danish and European legislation, the data from this manuscript cannot be shared as it contains information, which can lead to identification of individual participants.

## Author contributions

All authors contributed to the design of the study. Ms. Rydahl and Dr. Brund conducted the analyses. The manuscript was drafted by Ms. Rydahl and Dr. Østergaard, and was critically revised for important intellectual content by the remaining authors. The final version of the manuscript was approved by all authors prior to submission. Ms. Rydahl and Dr. Brund had full access to all of the data in the study and take responsibility for the integrity of the data and the accuracy of the data analysis.

## Acknowledgement

We thank the staff from the two general practices (Lægehuset i Vorgod and Lægerne Borggade) and from the outpatient units for unipolar depression and bipolar disorder for their contribution to recruitment of participants. Also, we are grateful to the participating patients.

## References

Aalbers, G., McNally, R.J., Heeren, A., de Wit, S., Fried, E.I., 2019. Social media and depression symptoms: A network perspective. J. Exp. Psychol. Gen. 148, 1454–1462. https://doi.org/10.1037/xge0000528

Beck, A.T., 2008. The evolution of the cognitive model of depression and its neurobiological correlates. Am. J. Psychiatry 165, 969–977. https://doi.org/10.1176/appi.ajp.2008.08050721

Berry, N., Emsley, R., Lobban, F., Bucci, S., 2018. Social media and its relationship with mood, self-esteem and paranoia in psychosis. Acta Psychiatr. Scand. https://doi.org/10.1111/acps.12953

Boase, J., Ling, R., 2013. Measuring Mobile Phone Use: Self-Report Versus Log Data. J. Comput. Commun. 18, 508–519. https://doi.org/10.1111/jcc4.12021

Boers, E., Afzali, M.H., Newton, N., Conrod, P., 2019. Association of Screen Time and Depression in Adolescence. JAMA Pediatr. 173, 853–859. https://doi.org/10.1001/jamapediatrics.2019.1759

Chou, H.T.G., Edge, N., 2012. “They are happier and having better lives than I am”: The impact of using facebook on perceptions of others’ lives. Cyberpsychology, Behav. Soc. Netw. 15, 117–121. https://doi.org/10.1089/cyber.2011.0324

Clement, J., 2019. Daily time spent on social networking by internet users worldwide from 2012 to 2018 (in minutes). URL https://www.statista.com/statistics/433871/daily-social-media-usage-worldwide/

Correll, C.U., Hauser, M., Penzner, J.B., Auther, A.M., Kafantaris, V., Saito, E., Olvet, D., Carrión, R.E., Birmaher, B., Chang, K.D., Delbello, M.P., Singh, M.K., Pavuluri, M., Cornblatt, B.A., 2014. Type and duration of subsyndromal symptoms in youth with bipolar I disorder prior to their first manic episode. Bipolar Disord. https://doi.org/10.1111/bdi.12194

Chang, K.D., Delbello, M.P., Singh, M.K., Pavuluri, M., Cornblatt, B.A., 2014. Type and duration of subsyndromal symptoms in youth with bipolar I disorder prior to their first manic episode. Bipolar Disord. https://doi.org/10.1111/bdi.12194

Deng, T., Kanthawala, S., Meng, J., Peng, W., Kononova, A., Hao, Q., Zhang, Q., David, P., 2019. Measuring smartphone usage and task switching with log tracking and self-reports. Mob. Media Commun. 7, 3–23. https://doi.org/10.1177/2050157918761491

Epskamp, S., Cramer, A.O.J., Waldorp, L.J., Schmittmann, V.D., Borsboom, D., 2012. Qgraph: Network visualizations of relationships in psychometric data. J. Stat. Softw. https://doi.org/10.18637/jss.v048.i04

Fruchterman, T.M.J., Reingold, E.M., 1991. Graph drawing by force‐directed placement. Softw. Pract. Exp. https://doi.org/10.1002/spe.4380211102

Gerson, J., Plagnol, A.C., Corr, P.J., 2017. Passive and Active Facebook Use Measure (PAUM): Validation and relationship to the Reinforcement Sensitivity Theory. Pers. Individ. Dif. 117, 81–90. https://doi.org/10.1016/j.paid.2017.05.034

Hanna, E., Monique Ward, L., Seabrook, R.C., Jerald, M., Reed, L., Giaccardi, S., Lippman, J.R., 2017. Contributions of Social Comparison and Self-Objectification in Mediating Associations between Facebook Use and Emergent Adults’ Psychological Well-Being. Cyberpsychology, Behav. Soc. Netw. 20, 172–179. https://doi.org/10.1089/cyber.2016.0247

Jabben, N., Arts, B., Jongen, E.M.M., Smulders, F.T.Y., Van Os, J., Krabbendam, L., 2012. Cognitive processes and attitudes in bipolar disorder: A study into personality, dysfunctional attitudes and attention bias in patients with bipolar disorder and their relatives. J. Affect. Disord. https://doi.org/10.1016/j.jad.2012.04.022

Junco, R., 2013. Comparing actual and self-reported measures of Facebook use. Comput. Human Behav. 29, 626–631. https://doi.org/10.1016/j.chb.2012.11.007

Kærsgaard, S., Meluken, I., Kessing, L. V., Vinberg, M., Miskowiak, K.W., 2018. Increased sensitivity to positive social stimuli in monozygotic twins at risk of bipolar vs. unipolar disorder. J. Affect. Disord. https://doi.org/10.1016/j.jad.2018.02.055

Kilford, E.J., Foulkes, L., Potter, R., Collishaw, S., Thapar, A., Rice, F., 2015. Affective bias and current, past and future adolescent depression: A familial high risk study. J. Affect. Disord. 174, 265–271. https://doi.org/10.1016/j.jad.2014.11.046

Kopeykina, I., Kim, H.J., Khatun, T., Boland, J., Haeri, S., Cohen, L.J., Galynker, I.I., 2016. Hypersexuality and couple relationships in bipolar disorder: A review. J. Affect. Disord. 195, 1–14. https://doi.org/10.1016/j.jad.2016.01.035

Krumpal, I., 2013. Determinants of social desirability bias in sensitive surveys: a literature review. Qual. Quant. 47, 2025–2047. https://doi.org/10.1007/s11135-011-9640-9

Lam, D.H., Jones, S.H., Hayward, P., 2010. Cognitive Therapy for Bipolar Disorder: A Therapist’s Guide to Concepts, Methods and Practice: Second Edition. https://doi.org/10.1002/9780470970256

Lam, D.H., Watkins, E.R., Hayward, P., Bright, J., Wright, K., Kerr, N., Parr-Davis, G., Sham, P., 2003. A randomized controlled study of cognitive therapy for relapse prevention for bipolar affective disorder: Outcome of the first year. Arch. Gen. Psychiatry. https://doi.org/10.1001/archpsyc.60.2.145

Mabe, A.G., Forney, K.J., Keel, P.K., 2014. Do you “like” my photo? Facebook use maintains eating disorder risk. Int. J. Eat. Disord. https://doi.org/10.1002/eat.22254

McCrae, N., Gettings, S., Purssell, E., 2017. Social Media and Depressive Symptoms in Childhood and Adolescence: A Systematic Review. Adolesc. Res. Rev. 2, 315–330. https://doi.org/10.1007/s40894-017-0053-4

Nadkarni, A., Hofmann, S.G., 2012. SOCIAL COMPARISONS ON FACEBOOK Running head: SOCIAL COMPARISONS ON FACEBOOK. NIH Public Access 52, 243–249. https://doi.org/10.1016/j.paid.2011.11.007.

OECD, 2019. Society at a Glance. https://doi.org/10.1787/9789264106154-6-en

Ortiz-Ospina, E., 2019. Rise of social media. Our World Data. URL https://ourworldindata.org/rise-of-social-media

Østergaard, S.D., 2017. Taking Facebook at face value: why the use of social media may cause mental disorder. Acta Psychiatr. Scand. https://doi.org/10.1111/acps.12819

Pregibon, D., 1981. Logistic Regression Diagnostics. Ann. Stat. https://doi.org/10.1214/aos/1176345513

Reinares, M., Colom, F., Sánchez-Moreno, J., Torrent, C., Martínez-Arán, A., Comes, M., Goikolea, J.M., Benabarre, A., Salamero, M., Vieta, E., 2008. Impact of caregiver group psychoeducation on the course and outcome of bipolar patients in remission: A randomized controlled trial. Bipolar Disord. https://doi.org/10.1111/j.1399-5618.2008.00588.x

Riehm, K.E., Feder, K.A., Tormohlen, K.N., Crum, R.M., Young, A.S., Green, K.M., Pacek, L.R., La Flair, L.N., Mojtabai, R., 2019. Associations between Time Spent Using Social Media and Internalizing and Externalizing Problems among US Youth. JAMA Psychiatry 21205, 1–8. https://doi.org/10.1001/jamapsychiatry.2019.2325

Scharkow, M., 2016. The Accuracy of Self-Reported Internet Use—A Validation Study Using Client Log Data. Commun. Methods Meas. 10, 13–27. https://doi.org/10.1080/19312458.2015.1118446

Schönfelder, S., Langer, J., Schneider, E.E., Wessa, M., 2017. Mania risk is characterized by an aberrant optimistic update bias for positive life events. J. Affect. Disord. https://doi.org/10.1016/j.jad.2017.04.073

Scott, J., Paykel, E., Morriss, R., Bentall, R., Kinderman, P., Johnson, T., Hayhurst, R.A.H., 2006. Cognitive-behavioural therapy for severe and recurrent bipolar disorders: Randomised controlled trial. Br. J. Psychiatry. https://doi.org/10.1176/foc.5.1.64

Shakya, H.B., Christakis, N.A., 2017. Association of Facebook Use With Compromised Well-Being: A Longitudinal Study. Am. J. Epidemiol. 185, 203–211. https://doi.org/10.1093/aje/kww189

Shaw, A.M., Timpano, K.R., Tran, T.B., Joormann, J., 2015. Correlates of Facebook usage patterns: The relationship between passive Facebook use, social anxiety symptoms, and brooding. Comput. Human Behav. 48, 575–580. https://doi.org/10.1016/j.chb.2015.02.003

Soo, S.A., Zhang, Z.W., Khong, S.J.E., Low, J.E.W., Vamadevan, Alhabsyi, S.H.B.T., Chew, Q.H., Sum, M.Y., Sengupta, S., Vieta, E., McIntyre, R.S., Sim, K., 2018. Randomized controlled trials of psychoeducation modalities in the management of bipolar disorder: A systematic review. J. Clin. Psychiatry. https://doi.org/10.4088/JCP.17r11750

Thorisdottir, I.E., Sigurvinsdottir, R., Asgeirsdottir, B.B., Allegrante, J.P., Sigfusdottir, I.D., 2019. Active and Passive Social Media Use and Symptoms of Anxiety and Depressed Mood Among Icelandic Adolescents. Cyberpsychol. Behav. Soc. Netw. 22, 535–542. https://doi.org/10.1089/cyber.2019.0079

Tromholt, M., 2016. The Facebook Experiment: Quitting Facebook Leads to Higher Levels of Well-Being. Cyberpsychology, Behav. Soc. Netw. 19, 661–666. https://doi.org/10.1089/cyber.2016.0259

Tursi, M.F.D.S., Baes, C.V.W., Camacho, F.R.D.B., Tofoli, S.M.D.C., Juruena, M.F., 2013. Effectiveness of psychoeducation for depression: A systematic review. Aust. N. Z. J. Psychiatry. https://doi.org/10.1177/0004867413491154

Vannucci, A., Flannery, K.M., Ohannessian, C.M.C., 2017. Social media use and anxiety in emerging adults. J. Affect. Disord. 207, 163–166. https://doi.org/10.1016/j.jad.2016.08.040

Verduyn, P., Lee, D.S., Park, J., Shablack, H., Orvell, A., Bayer, J., Ybarra, O., Jonides, J., Kross, E., 2015. Passive Facebook usage undermines affective well-being: Experimental and longitudinal evidence. J. Exp. Psychol. Gen. 144, 480–488. https://doi.org/10.1037/xge0000057

Wang, C., Shao, X., Jia, Y., Ho, R.C., Harris, K.M., Wang, W., 2020. Peripherally Physiological Responses to External Emotions and Their Transitions in Bipolar I Disorder With and Without Hypersexuality. Arch. Sex. Behav. https://doi.org/10.1007/s10508-019-01615-8

Yoon, S., Kleinman, M., Mertz, J., Brannick, M., 2019. Is social network site usage related to depression? A meta-analysis of Facebook–depression relations. J. Affect. Disord. 248, 65–72. https://doi.org/10.1016/j.jad.2019.01.026

